# Development and formative application of the Health Data Readiness Level framework for federated health-data services

**DOI:** 10.64898/2026.07.23.26358713

**Authors:** David Seymour, Roger Halliday, Jon Smart, Frances Burns

## Abstract

**Objectives:** To develop a multidimensional framework for assessing organisational and system readiness for federated health-data services, and to report its formative application across three heterogeneous UK ecosystems.

**Methods:** The Health Data Readiness Level (HDRL) framework was developed from a structured landscape review of 56 maturity and readiness frameworks, first-principles requirements analysis, artificial-intelligence-assisted synthesis with human source verification, and stakeholder refinement. It comprises 64 indicators in eight domains and five ordered levels. Formative application examined three heterogeneous UK health-data research ecosystems using documentary evidence, professional stakeholder input, workshops, a structured right-of-reply process, cross-case calibration, and two illustrative research use cases. Analysis was descriptive; the application was not designed as psychometric validation or a league table.

**Results:** All 64 indicators were scoreable in each case. Publicly reported profiles ranged from Developing (Level 2–3) to Managed (Level 3–4); all three cases met the proposed minimum for five foundational indicators. Recurring constraints concerned evidence of measured service performance, national-scale primary-care data access, cross-jurisdiction governance reciprocity, workforce capacity, and sustainable funding. Pandemic-era four-nation research was delivered through coordinated local analyses and meta-analysis, rather than routine automated federation.

**Discussion:** HDRL operationalises a broad service- and system-readiness view that complements technical and governance specifications. Content validity, inter-rater reliability, aggregation choices, responsiveness and predictive validity remain to be established.

**Conclusion:** HDRL is an evidence-informed candidate improvement and planning instrument for federated health-data services. It should not yet be used as an accreditation standard or as an official participation threshold for the UK’s Health Data Research Service.

**Key messages:** *What is already known on this topic:*

- Readiness for services supporting federated health-data research depends on governance, data, service operations, semantics, workforce, sustainability, infrastructure and public legitimacy; existing frameworks and blueprints address important but partly separate components.

*What this study adds:*

- HDRL operationalises this broader system and service perspective through 64 indicators, eight domains and five ordered maturity levels, and reports a public-safe formative application across three heterogeneous UK ecosystems.

*How this study might affect research, practice or policy:*

- HDRL can support structured improvement and investment planning, but mapping to related models and independent testing of content validity, scoring reliability and outcomes are needed before any accreditation or participation threshold use.

**Plain-language summary:** Health-data research increasingly depends on several secure services working together without moving sensitive records into one central location. Readiness for this kind of federation depends on more than technology: services also need suitable data, consistent meanings, lawful and efficient access processes, public legitimacy, sustainable funding, and enough skilled staff. HDRL brings these conditions together in 64 indicators across eight domains and five maturity levels. Its formative application across three different UK operating models showed that it can structure evidence, surface dependencies, and focus improvement planning. It also highlighted gaps in measurable service performance, primary-care data access, coordination of approvals, workforce capacity and funding. The framework remains at an early stage of validation. It is best used to support improvement and investment planning, not as accreditation or an official threshold for participation in the UK Health Data Research Service.

## Introduction

Health systems are expected to support research, innovation, service planning, and regulatory science using routinely collected data while protecting privacy, maintaining public trust, and preserving accountable local stewardship. In the United Kingdom, this ambition is being pursued through the Health Data Research Service (HDRS), backed by up to £600 million from the UK Government and Wellcome and intended to simplify secure access to national-scale health data for approved research.[1-3] Reviews of the UK landscape have nevertheless described fragmented access routes, uneven data availability, slow approvals, and insufficiently coordinated infrastructure.[4-6]

Federated health-data research is a sociotechnical undertaking: participating nodes must align governance, data semantics, service operations, security, workforce, financing, and public legitimacy. Readiness therefore differs from the mere presence of a trusted research environment (TRE) or secure data environment. It refers to the evidenced ability of an organisation, service, or system to contribute reliably to a defined federated operating model.

Maturity models can support baseline assessment and improvement planning, but their credibility depends on transparent development methods, coherent level definitions, stakeholder relevance, and evaluation of reliability and validity.[7-10] Existing instruments address important components. FAIR and the Research Data Alliance FAIR Data Maturity Model concern data stewardship and reusability.[11,12] Five Safes provides a governance lens for safe use.[13] UK TRE guidance, the Standardised Architecture for Trusted Research Environments (SATRE), and the DARE UK Federated Architecture Blueprint specify technical and operational capabilities.[14-16] The Organisation for Economic Co-operation and Development provides health-data governance principles.[17]

Luong and colleagues recently proposed a six-domain maturity-model framework for federated TRE networks, mapped to SATRE and the DARE UK blueprint, as a foundation for further specification and validation.[18] HDRL Framework’s distinct contribution is the operationalisation of a broader service- and system-readiness scope across data coverage, semantics, governance, research delivery, public trust, sustainability, workforce and infrastructure.

This study had two objectives: to describe the development of HDRL and to report a formative field application across three heterogeneous UK health-data research ecosystems. The work tested practical usability and generated improvement priorities. It did not establish psychometric validity, accredit participating services or rank nations.

## Methods

### Study design and setting

HDRL was developed and applied between December 2025 and April 2026 as part of a commissioned organisational readiness assessment. Three cases represented deliberately different operating arrangements: a networked national ecosystem, a national data service, and an integrated service/platform arrangement. The cases were treated as formative applications rather than exchangeable units in a comparative league table.

The assessment related to potential participation in the emerging HDRS. The framework and the five proposed foundational indicators are project outputs; they are not official HDRS requirements.

### Framework development

Development comprised four linked activities. First, a structured AI-supported landscape review considered 53 maturity, readiness, capability, governance, data-quality, digital-government, research-infrastructure, and workforce frameworks. A further three design sources were drawn directly from prior experience by the lead author: the B1MG Maturity Level Model,[19] SATRE,[15] and Building Trusted Research Environments,[14] giving 56 frameworks in total. Extraction considered purpose, architecture, domains, measurement approach, validation evidence, strengths, limitations, and relevance to health-data research. It was not a systematic review, and the work was not prospectively registered. A complete reproducible search appendix is not available. The 56 frameworks reviewed, with type and relevance to health-data research, are listed in Supplementary Table S3.

Second, AI-enabled first-principles deep research and analysis identified necessary, enabling, and excellence conditions for federated health-data research across patients and the public, researchers, health services, data controllers, regulators, funders, industry, service operators, and policy makers. This analysis helped distinguish baseline requirements from capability-specific and optional indicators.

Third, Claude, Gemini and ChatGPT were used as structured synthesis and review aids. Each system received a defined task specification, and outputs were compared for agreement, omissions and contradictions. The specific frontier AI models used, and additional information are listed in Supplementary Table S4.

Fourth, an Oversight Group comprising senior representatives of the three assessed ecosystems reviewed version 0.9. Their feedback refined definitions, applicability classes, level descriptors, and the proposed foundational set. This was stakeholder refinement, not independent content-validity testing.

### Framework architecture

Version 1.0 contains 64 indicators: 43 Core and 21 Enhancement indicators. Indicators are organised into eight domains and five ordered maturity levels: Level 1 Initial, Level 2 Developing, Level 3 Defined, Level 4 Managed, and Level 5 Optimising. Core indicators include baseline, capability-specific, and outcome/context classes. Outcome/context indicators are descriptive and are not treated as prerequisites.

Five indicators were proposed as foundational for baseline participation, each with a project-defined minimum of Level 3: legal basis for research processing; a data access committee; statistical disclosure control; security certification and audit; and security operations. Level 4 was designed to require measured and standardised performance supported by published or otherwise auditable evidence. This evidence rule was intended to guard against over-scoring asserted capability, but it also creates a methodological tension between capability maturity and evidence visibility.

### Field application and evidence

Evidence was gathered through 16 stakeholder interviews and follow-up discussions across the three nations (23 participants: Scotland 5, Wales 8, Northern Ireland 10), spanning senior strategic leadership, data services and technical leads, governance and legal specialists, programme management and policy advisers, conducted between December 2025 and February 2026; three national workshops (31 participants: Scotland 18, Wales 6, Northern Ireland 7); and a joint three-nations synthesis session. Stakeholders were informed that meetings would be recorded and transcribed for note preparation and that the write-up would be returned for factual checking. Recordings were transcribed and a configured Gemini Gem in a managed Google Workspace account prepared AI-assisted notes; after each interview these were shared with the stakeholder(s), who could correct misinterpretations or mistakes and object to their transfer to Research Data Scotland as a project artefact. ChatGPT and Claude were used as additional scoring reviewers alongside the primary assessor; every indicator score and proposed right-of-reply change was reviewed by a human, who made or approved the final decision.

One primary assessor assigned indicator scores using an evidence hierarchy: uncorroborated testimony could support no more than Level 2; formal documentation could support Level 3; measured performance and external assurance could support Level 4; and systematic improvement with benchmarking was required for Level 5.

Each case received a structured right of reply to correct factual errors and submit further evidence against specific indicators. Proposed changes were adjudicated against the same descriptors and then reviewed across cases for consistency. The organisations assessed were represented among the co-authors but were not directly involved in making the assessments; right of reply and cross-case calibration improved factual accuracy but do not remove the risk of bias.

### Illustrative use cases

Two use cases were used to probe whether framework findings aligned with practical delivery dependencies. The retrospective COALESCE study analysed severe COVID-19 outcomes across the four UK nations.[20] A prospective shingles-vaccine and dementia-risk study announced by GSK, the UK Dementia Research Institute, and Health Data Research UK was used to examine routine-state requirements for longitudinal primary-care and outcome data.[21] These cases were illustrative stress tests, not criterion or predictive validation of HDRL.

### Analysis and public-reporting boundary

Analysis was descriptive. We examined whether all indicators could be scored, the distribution of maturity levels, proposed foundational-indicator status, recurring evidence gaps, and operational dependencies identified across the cases. Because maturity levels are ordinal and the assessment units were heterogeneous, the revised analysis does not report arithmetic means or country domain medians.

Country-specific reporting in this article is deliberately limited to aggregate profiles and score distributions explicitly published in the Final Report.[26] The unpublished indicator-by-country matrix, country domain-median heatmap, underlying evidence records, and right-of-reply material are excluded.

### Patient and public involvement

The framework includes public trust and lay governance as assessment domains, but the development study did not include a dedicated independent patient and public co-design or content-validity panel. This is a priority for the next validation phase.

## Results

### Framework output and usability

The resulting framework covered eight domains (Table 1) and retained a common five-level architecture across all indicators. All 64 indicators could be applied to each of the three cases, indicating practical coverage across different organisational forms. This finding supports usability, but not reliability or validity.

**Table 1.** HDRL framework architecture.

| Ref | Domain | Indicators | Primary focus |
| --- | --- | --- | --- |
| A | Data Coverage and Federation | 10 (7 Core, 3 Enhancement) | Data availability, linkage, federation, and extended data modalities |
| B | Data Semantics and Quality | 6 (4 Core, 2 Enhancement) | Common models, terminology, quality monitoring, and metadata |
| C | Governance and Access | 11 (9 Core, 2 Enhancement) | Legal basis, access processes, cross-border coordination, and assurance |
| D | Research Integration and Market | 8 (5 Core, 3 Enhancement) | Research users, support, multi-site work, commercial access, and trials |
| E | Public Trust and Transparency | 7 (6 Core, 1 Enhancement) | Transparency, lay governance, engagement, public benefit, and legitimacy |
| F | Sustainability | 6 (2 Core, 4 Enhancement) | Funding horizon, pricing, financial resilience, and value demonstration |
| G | Workforce and Culture | 7 (4 Core, 3 Enhancement) | Capacity, professional roles, skills, service culture, and collaboration |
| H | Infrastructure and Compute | 9 (6 Core, 3 Enhancement) | TRE architecture, user environment, compute, security, and responsible AI |

### Publicly reportable readiness profiles

The aggregate findings approved for public release are shown in Table 2. The same distributions are shown graphically in Figure 1.

**Table 2.** Publicly reportable aggregate readiness profiles.

| Ecosystem | Public profile and status | Score distribution | Proposed foundational indicators |
| --- | --- | --- | --- |
| Wales / SAIL Databank | Managed (L3-L4)<br>Public report status: READY | L1=0; L2=2; L3=43; L4=16; L5=3; L4+=19 | All 5 ≥ L3; 4 at L4 |
| Scotland / Scottish Safe Haven Network | Defined (L3)<br>Public report status: READY BUT GP DATA GAP | L1=0; L2=10; L3=48; L4=6; L5=0; L4+=6 | All 5 ≥ L3; 1 at L4 |
| Northern Ireland / NITRE HSC Data Institute | Developing (L2-L3)<br>Public report status: READY WITH CONDITIONS | L1=5; L2=27; L3=31; L4=1; L5=0; L4+=1 | All 5 at L3 |

**Note:** Source: *Three Nations Readiness Assessment—Final Report, July 2026* [26]. Foundational indicators and thresholds are project proposals, not official HDRS requirements. L4+ denotes indicators at Level 4 or Level 5.

**Figure 1.**
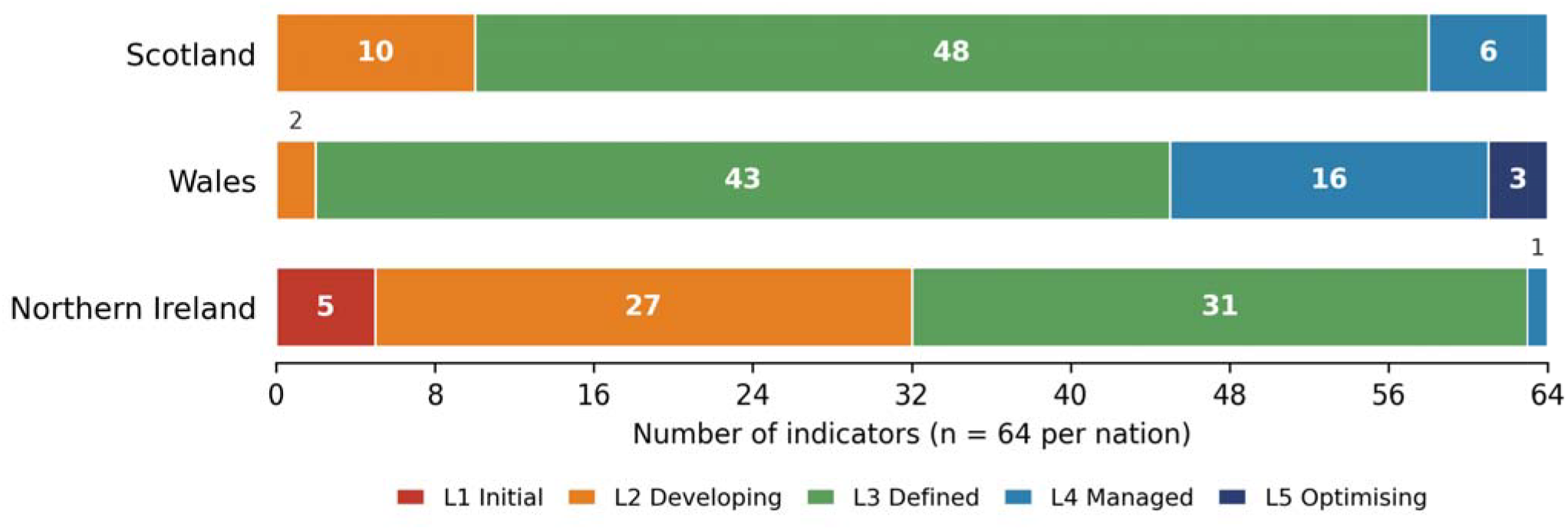
Distribution of HDRL indicator scores by nation after adjudication (n = 64 indicators per nation). Values as published in the project final report.[26] Alt text: *Horizontal stacked bar chart showing the distribution of 64 HDRL indicator scores for Scotland, Wales, and Northern Ireland across maturity Levels 1 to 5. Scotland: 10 indicators at Level 2, 48 at Level 3, and 6 at Level 4. Wales: 2 at Level 2, 43 at Level 3, 16 at Level 4, and 3 at Level 5. Northern Ireland: 5 at Level 1, 27 at Level 2, 31 at Level 3, and 1 at Level 4*.

Profiles ranged from Developing-to-Defined to Managed. All three cases met the project-defined Level 3 minimum for the five proposed foundational indicators. These labels describe evidence against HDRL version 1.0 and must not be interpreted as official HDRS accreditation or participation decisions.

### Cross-case findings

A recurring finding was an evidence-assurance gap: services could often demonstrate that a process or capability existed but had less consistent evidence of measured service performance. Common examples concerned time to data, data latency, linkage quality, metadata completeness, user experience, workforce resilience, and service-level reporting. This finding is qualitative. Post hoc keyword count of evidence-gap fields has not been included because it was not a prospectively specified or independently replicated analysis.

The application also distinguished capability from capacity. A service may possess the technical and governance capability to deliver a type of research but lack sufficient analyst, information-governance, or operational capacity to deliver it routinely at greater scale. Across cases, national-scale primary-care data access, reciprocal or reusable governance processes, workforce capacity, and sustainable funding emerged as important dependencies. Wales’s approximately 86% GP population coverage through its WLGP dataset illustrates the upper end of the primary care data access dependency; Scotland’s GP data at time of assessment reached only around 17% of the population, available at regional scale for Lothian only through DataLoch.

The two use cases reinforced these dependencies. COALESCE achieved four-nation population analysis through separate analyses in national secure environments followed by meta-analysis; it did not demonstrate routine automated federation of person-level data.[20] The prospective use case highlighted the importance of longitudinal primary-care data and the cumulative effect of multiple approval pathways. No inference was made about the internal status of the announced study.

## Discussion

### Principal findings

HDRL provides a structured way to examine readiness for federated health-data research across dimensions that are often assessed separately. Its formative application showed that a common indicator set could be used across three different ecosystem arrangements and could identify concrete improvement priorities. The most consistent signal was not simply the absence of capability, but a gap between capability claims and comparable evidence of operational performance.

The current evidence supports describing HDRL as an evidence-informed candidate maturity and improvement framework. It does not claim to be scientifically validated, nor that a given level guarantees speed or predictability, or that the proposed foundational indicators are non-negotiable conditions for HDRS participation.

### Relationship to existing work

HDRL complements rather than replaces FAIR, Five Safes, SATRE, and the DARE UK blueprint.[11-16] Those resources provide principles, data-practice criteria, and technical or architectural requirements. HDRL adds a service- and system-readiness lens incorporating data availability, public legitimacy, financing, workforce, and research delivery. SATRE version 2.0, published in July 2026 after this assessment concluded, adds a fifth pillar covering federation between trusted research environments and raises selected public involvement and regulatory compliance statements from recommended to mandatory.[15] The application reported here used SATRE version 1.0, and the HDRL mapping to SATRE has not yet been revised against version 2.0.

Luong and colleagues propose a six-domain maturity-model framework for federated TRE networks and call for indicators and maturity descriptors to be added, refined and validated.[18] The overlap with HDRL in governance, operations and infrastructure is material, but the instruments are not interchangeable. HDRL is more granular and extends the unit of analysis across data coverage, semantics, research delivery, public trust, sustainability and workforce. The novelty of HDRL is therefore operationalisation and formative application of this broader readiness instrument. Future work should map the 64 HDRL indicators explicitly to the Luong framework, the Federated Architecture Blueprint, and the SATRE version 2.0 federation pillar, extending the initial mapping to SATRE version 1.0 undertaken as part of the HDRL framework development.

### Evidence maturity and scoring design

Requiring evidence prevents unsupported self-assessment from inflating scores and creates a useful improvement agenda around transparency and assurance. However, an organisation with strong internal performance but limited public reporting may receive the same score as an organisation with weaker capability. A future version should record at least two dimensions: capability maturity and confidence or visibility of evidence. External publication of metrics may then inform the evidence-confidence rating without being treated as the capability itself.

The five maturity levels are ordinal categories, not equal numerical intervals. Counts and profiles are therefore more defensible than means, fine-grained rankings, or untested weighted totals. The rationale for indicator weights, any domain aggregation, and the foundational set requires sensitivity analysis and independent stakeholder testing.

### Implications for federated services

A central data discovery portal does not by itself create a single operational pathway. Federation also requires compatible metadata, reusable approvals and agreements, predictable service performance, distributed operational support, and capacity at each node. Recent commentary has argued that HDRS can deliver major public benefit only if it is grounded in real-world care delivery and connected internationally, rather than built as a stand-alone centralised system — a concern this paper’s federation findings substantiate empirically.[22]

The use cases suggest a pragmatic sequence: begin with harmonised specifications and coordinated analyses in existing secure environments, then automate components where governance, semantics, and infrastructure are sufficiently mature.

Public trust is a substantive part of readiness. Previous UK experience shows that legal authority and technical safeguards do not by themselves secure social legitimacy.[23-25] Future validation should therefore include public contributors directly in defining evidence standards, especially for transparency, commercial access, public benefit, and opt-out or objection mechanisms.

### Strengths and limitations

Strengths include an intentionally multidimensional scope, use of real organisational evidence, a structured evidence hierarchy, opportunities for factual correction, and application across heterogeneous cases. The project also made explicit that artificial intelligence supported deep research to develop the framework, note preparation, synthesis and scoring review, while a human reviewed every score and proposed right-of-reply change and remained responsible for all final decisions.

The study was commissioned as a consulting project, not research and so there are consequent limitations. The landscape review is not reproducible to systematic-review standards. Indicator content was refined by a small, interested Oversight Group rather than an independent multidisciplinary panel. A single primary human assessor scored all cases, so human inter-rater reliability is unknown and whilst the deployed AI models each scored the same evidence, the approach was one of adversarial challenge to the lead assessor.

The units of assessment differed given the heterogeneous nature of the health data research services across the devolved nations and the assessed organisations were represented among the authors. No construct, criterion, or predictive validity was tested. The proposed foundational indicators and Level 4 evidence threshold

are judgement-based. Aggregation and weighting were not subjected to sensitivity analysis. The detailed score matrix is management information specific to each service and not intended to be in the public domain, limiting independent reproduction of country-specific analyses.

The study lacked a dedicated patient and public co-design panel and was conducted within one UK policy setting across three national jurisdictions. International transferability remains untested. Readiness changes over time, so any assessment should be versioned and dated.

### Validation programme

Before HDRL is used for accreditation, funding allocation, or formal participation decisions, a proportionate validation programme, in priority order, would include: a reproducible review appendix and complete source-framework list; independent expert and public content-validity assessment, using a Delphi process or content-validity indices; duplicate blinded scoring of at least one complete case or a substantial stratified indicator sample with percentage agreement and weighted kappa; sensitivity analysis of weighting, aggregation, foundational thresholds, and missing evidence; separation of capability and evidence-confidence ratings; and prospective reassessment to test responsiveness and whether scores predict delivery outcomes.

This bar is deliberately conservative; established accreditation regimes in this field typically rest on documented procedure, audit and periodic review rather than psychometric validation.

The framework is published openly under CC BY 4.0. Its website provides the full indicator set and maturity descriptors, methodology and usage guidance, the applied v1 method source files, a plain-language glossary, and citation and responsible-reuse guidance, using evidence-informed and formative language [27].

## Conclusion

HDRL addresses an important gap between high-level governance principles and technical specifications by assessing the broader conditions required for providing federated health-data research services. The three-case application supports practical usability and identifies recurring system dependencies. It does not yet establish reliability, validity, or accreditation fitness. HDRL should therefore be used as a structured improvement and investment-planning instrument while an independent validation programme is completed.

## Supporting information

Supplementary_Table_S2_HDRL_Aggregate_Results

Supplementary_Table_S4_HDRL_AI_Tool_Use

Supplementary_Table_S1 HDRL Indicator Catalogue

Supplementary_Table_S3_HDRL_Source_Frameworks

## Ethics and governance

Research Data Scotland commissioned this work as a consultancy-based organisational readiness assessment; hence NHS Research Ethics Committee review was not required. Stakeholders contributed in their professional capacities; no patients or service users were recruited, and no patient-level data were used. Recordings and transcripts were stored on an encrypted MacBook and associated iCloud storage for the duration of the project and deleted at project end. Curated interview notes, workshop write-ups and Oversight Group minutes were retained. Interview recordings and transcripts were processed only within a managed Google Workspace account not through consumer-tier AI tools.

## Data availability

The public framework and public indicator catalogue are available through the HDRL website [27] and Supplementary Table S1. Public aggregate assessment results are available in the Final Report [26] and Supplementary Table S2. The indicator-by-country matrix, underlying restricted evidence records and right-of-reply material are not available because they are management information and contain assessment detail not intended for publication. The descriptive results in Table 2 can be Sreproduced from Supplementary Table S2. The authorised Final Report and its public download are available from the Research Data Scotland publication page cited in reference 26. The 56 frameworks considered during development are listed in Supplementary Table S3, whilst the AI model use is specified in Supplementary Table S4.

## Artificial intelligence transparency statement

Claude (Anthropic; Opus 4.5, Opus 4.6, Opus 4.7, Opus 4.8, Sonnet 5, Fable 5), Gemini (Google; Gemini 3 Pro, Gemini 3.1 Pro, accessed via a Google Workspace Business Plus account with the AI Ultra Access add-on, since retired), and ChatGPT (OpenAI; GPT-5.2 Pro, GPT-5.4 Pro, GPT-5.6 Sol Pro, all on a ChatGPT Pro subscription) were used across framework development, field application, scoring review, and manuscript drafting between December 2025 and July 2026, with the specific model generation varying by project phase as detailed in Supplementary Table S4. The Google Workspace account’s enterprise terms exclude customer data from model training by default; for Claude Pro and ChatGPT Pro, controls permitting user data to be used for model training were disabled by user action. All AI outputs were reviewed by a human, who made or approved every final score, interpretation and publication decision.

## Author contributions

David Seymour: Conceptualization, Methodology, Investigation, Data curation, Formal analysis, Project administration, Writing—original draft, Writing—review and editing. Roger Halliday: Funding acquisition, Resources, Investigation, Project administration, Writing—review and editing. Jon Smart: Resources, Investigation, Writing—review and editing. Frances Burns: Resources, Investigation, Writing—review and editing. All authors: final approval and accountability.

## Acknowledgements

The authors thank the professional stakeholders who contributed evidence, interviews, workshops, right-of-reply review and synthesis discussions.

